# The autonomic age gap: a machine learning approach to assess biological-calendar age deviations

**DOI:** 10.1101/2025.04.16.25325919

**Authors:** Andy Schumann, Yubraj Gupta, Denis Gerstorf, Ilja Demuth, Maja Olecka, Christian Gaser, Karl-Jürgen Bär

## Abstract

Machine learning has emerged as a valuable tool in precision medicine and aging research. Here, we introduce the *autonomic age gap*, a novel metric quantifying the individual deviation between machine-learning–estimated biological age and chronological age, based on autonomic nervous system function. We collected high-resolution electrocardiograms and continuous blood pressure recordings at rest from 1,012 healthy individuals. From these signals, 29 autonomic indices were extracted, encompassing time-, frequency-, and symbol-domain heart rate variability, cardiovascular coupling, pulse wave dynamics, and QT interval features. Based on those parameters, a Gaussian process regression model was trained on 879 participants to estimate chronological age referred to as *autonomic age*. The model was used to estimate the deviation from expected healthy aging, the *autonomic age gap*, in an independent validation set and two test sets stratified by cardiovascular risk (CVR) using the Framingham. The was evaluated via the autonomic age gap.

High CVR individuals had a significantly increased *autonomic age gap* of 9.7 years compared to the low CVR group and the validation set. In contrast, the low CVR group had a negative age gap of -2.2 years on average. Predictions in the validation sample closely matched calendar age with a deviation below 0.5 years. Additionally, in the high-risk group, the slope of predicted versus actual age suggested accelerated physiological aging.

These findings highlight the autonomic age gap as a sensitive and interpretable marker of cardiovascular risk and aging, offering potential clinical utility for early risk detection and longitudinal health monitoring.

## Introduction

Machine learning has emerged as a transformative tool in the study of biomarkers of the aging process, offering insights with significant diagnostic, prognostic, and predictive relevance in age-related diseases (1). By leveraging large datasets, machine learning algorithms can identify patterns and relationships among biological markers—such as epigenetic modifications, proteomic profiles, organ structural changes, or metabolic changes—that are closely associated with aging and disease progression (2–4). These biomarkers can aid in early diagnosis, risk stratification, and monitoring of conditions like cardiovascular disease, neurodegenerative disorders, and cancer (5, 6).

Machine learning can be applied to track age-related changes in specific organ systems, helping to characterize normal, healthy aging processes. If an individual’s estimated age, derived from a machine learning model of healthy aging trained on large datasets, is larger than their actual calendar age, then this can serve as a risk marker. Such an age equivalent or risk age may enhance communication with patients: By presenting risk in a relatable way and straightforward manner, it may improve adherence to recommended lifestyle modifications and pharmacotherapies (7).

To assess neurological health, the concept of brain age has become widely recognized. The biological state of the brain can deviate significantly from the normal due to factors like neurodegenerative processes (8). A positive brain-age gap, where the brain appears older than the chronological age, signals accelerated aging and may indicate a heightened risk of conditions such as Alzheimer’s and Parkinson’s diseases. In contrast, a negative gap suggests delayed aging and potentially better cognitive maintenance and resilience. Brain age prediction models, often based on MRI scans, provide valuable insights for detecting and monitoring neurodegenerative disorders, aiding early intervention, and advancing neurological research (9). In contrast, the application of machine learning to cardiovascular data remains less explored, despite the critical role of cardiovascular health in aging. Although numerous age-related changes in cardiovascular systems have been described, their integration into predictive models for biological age or disease risk lags behind neurological research.

Increased arterial and ventricular stiffness, reduced myocardial contractility, impaired beta-adrenergic and parasympathetic function, and degenerative changes in the conduction system are commonly observed in aging populations. Factors such as endothelial dysfunction and oxidative stress contribute to arterial stiffening, reduced compliance, and diminished absorption of arterial pulse waves. These changes promote conditions like sustained hypertension, atherosclerosis, and thrombosis (10). Indicators of such age-related changes include broad pulse waves, elevated pulse wave velocity, and high systolic blood pressure. Structural alterations, such as sinoatrial pacemaker cell loss and left ventricular hypertrophy, alongside functional changes in autonomic reflex receptors (e.g., baroreceptors), further affect cardiovascular regulation in aging (11, 12).

Reduced vagal influence on the cardiovascular system, such as diminished respiratory sinus arrhythmia, leads to higher resting heart rates and lower heart rate variability (HRV) in older adults (13, 14). Resting heart rate and HRV also predict cardiovascular disease and overall mortality later in life.

Although some age equivalents appear to be quite common for risk prediction in a clinical context (15), the potential of machine learning has not been fully leveraged. Existing markers such as “Heart Age”, “Vascular Age”, “Arterial Age” or “Coronary Age” are based on cardiovascular risk factors that are rather easy to assess and used in clinical routine (16–20).

As a first step towards harnessing the potential of machine learning, we recently developed a model to estimate age based on cardiovascular function (21). In total, 29 cardiovascular indices were estimated as input features to the age-prediction model, including heart rate variability, blood pressure variability, baroreflex function, pulse wave dynamics, and QT interval characteristics. Four approaches were tested to estimate the calendar age of healthy individuals: relevance vector regression, Gaussian process regression (GPR), support vector regression, and linear regression. Through a five-fold cross-validation process, the GPR model demonstrated the best performance in estimating calendar age, achieving a high correlation and a mean absolute error of 5.6 years. Interestingly, the estimated age of obese individuals (body mass index above 30 kg/m²) was markedly higher by up to 6 years on average compared to normal-weight participants indicating an advanced cardiovascular aging of in this group.

However, body mass index (BMI) is only one factor influencing cardiovascular risk. For example, the Framingham multivariate risk score is a widely used tool for assessing 10-year atherosclerotic cardiovascular risk (16). Based on a cohort of 8,491 Framingham study participants, sex-specific risk functions were developed using age, BMI or cholesterol levels, systolic blood pressure, hypertension treatment, smoking, and diabetes status. Independent studies have validated these findings, demonstrating that BMI-based and laboratory-based risk scores provide comparable predictive accuracy for both fatal and nonfatal cardiovascular events (22, 23).

The current study explores whether the cardiovascular aging model reveals different age gaps for individuals with high or low cardiovascular risk. Using the Framingham algorithm, we estimated 10-year cardiovascular risk and compared two groups on age and sex: one with higher and one with lower risk than the age-dependent norm. The autonomic age gap was assessed as a deviation from healthy aging and compared between groups.

## Materials and methods

### Participants

We investigated data of healthy 1,012 individuals recorded in our laboratory. The majority of datasets have been made publicly available already (24). In addition, we assessed another sample of 132 healthy individuals. None of the subjects had any history of a neurological or psychiatric disorder. Exclusion criteria were any medical conditions, illegal drugs or medication potentially influencing cardiovascular function. Thorough physical examination, resting electrocardiography (ECG) and routine laboratory parameters (electrolytes, basic metabolic panel, and blood count) had to be without any pathological finding. All participants provided written informed consent prior to participating in the study. The study protocol was approved by the Ethics Committee of the University Hospital of Jena.

### Cardiovascular risk estimation

We used the updated Framingham risk calculation model to predict the 10-year risk of atherosclerotic cardiovascular disease, including coronary heart disease, stroke, peripheral vascular disease, and heart failure (D’Agostino et al., 2008). Cardiovascular risk (CVR) was estimated based on age, sex, systolic blood pressure (SBP), BMI, hypertension or diabetes diagnosis, smoking status, and current use of antihypertensive medications. The calculation algorithm is publicly available at the Framingham Heart Study website (https://www.framinghamheartstudy.org/fhs-risk-functions/cardiovascular-disease-10-year-risk/). CVR estimates range from 0.3% to over 30%, with a practical upper limit of 98.5%. Additionally, normal risk at a given age can be approximated by assuming an SBP of 125 mmHg and a BMI of 22.5 kg/m².

### Training and testing samples

From the sample of 1,012 individuals, we assessed CVR in 199 participants aged 30 to 74 years, for whom complete data were available to calculate the Framingham risk score, including sex, age, systolic blood pressure, smoking status, hypertensive treatment, BMI, and diabetes diagnosis. From this dataset, we selected a high CVR group of 72 participants (31 men, 41 women, mean age 53.1 ± 12.9 years) whose risk was at least 0.5% above the normal population, and a low CVR group of 61 participants (20 men, 41 women, mean age 52.5 ± 13.5 years) with a risk at least 0.5% below the normal population. From the remaining 879 individuals (351 men, 528 women, mean age 31.8 ± 14 years), 70 were randomly selected for validation to assess model performance, while the remaining 809 individuals (324 men, 485 women, 31.8 ± 13.9 years) were used for training the model.

### Data acquisition and feature extraction

Continuous non-invasive blood pressure and electrocardiogram (ECG) were acquired simultaneously over 20 min using either a Task Force Monitor® (TFM, CNSystems Medizintechnik GmbH, Graz, Austria) or MP150 (BIOPAC Systems Inc, Goleta, CA, USA). Participants were in a supine position under well-controlled resting conditions in our laboratory. The assessment took place in a completely quiet and fully shaded room, with a constant illumination level. Following usual practice, the first five minutes were excluded from the analysis.

From these signals, we estimated 29 cardiovascular features including time, frequency and symbol domain heart rate variability, cardiovascular parameters, pulse wave dynamics and QT interval characteristics (see 21). Mean heart rate (HR), root mean square of successive BBI (RMSSD), standard deviation of BBI (SDNN), low to high frequency power ratio (25), deceleration capacity (26), Renyi entropy with base ¼ (27), sample entropy (28), compression entropy (29) and forbidden words (30) were derived from BBI time series. Mean and standard deviation of corrected QT intervals (31) and the QT variability index (QTvi) were estimated (32). Mean and standard deviation of systolic (SBP) and diastolic blood pressure values (DBP) per heart beat interval were extracted (33). Pulse pressure was calculated as difference between SBP and DBP. Using the dual sequence method, baroreflex sensitivity was calculated as marker of cardic changes due to blood pressure alterations (34). Mean values and standard deviation of the pulse transit time, pulse rise time, pulse wave duration, pulse wave velocity and time delay of the dicrotic notch were estimated on blood pressure signals (35).

### Autonomic age estimation

We applied machine learning in order to estimate effects of healthy cardiovascular aging based on 29 cardiovascular autonomic indices. The autonomic age gap represents the deviation between the estimated age from this model and people’s actual chronological age. A positive age gap indicates advanced aging relative to one’s chronological age, whereas a negative gap suggests delayed aging processes. A Gaussian process regression model with a squared exponential kernel and a constant basis function was applied (21). We corrected the model output for a potential linear dependency of the estimated on the calendar age (age bias) by fitting a linear regression model in the validation sample (36). The age predictions in the high CVR and low CVR groups were corrected for that bias using the estimated slope and intercept (37).

### Feature importance

To identify the most influential cardiovascular features contributing to the age predictions, we employed the SHapley Additive exPlanation (SHAP) algorithm (38). SHAP analysis was performed in the validation set of healthy controls randomly selected from the training data. Absolute SHAP values were extracted using a KernelExplainer and averaged to evaluate importance of each input feature.

### Statistical analysis

The accuracy of age estimation was assessed in the validation sample using mean absolute error (MAE) and root mean squared error (RMSE). Differences between estimated age and actual calendar age were compared between healthy participants with low and high cardiovascular risk (CVR) using two-sample t-tests. Individual risk factor comparisons were conducted using an independent t-test for continuous variables and a Wald test for dichotomous variables.

## Results

The final sample under investigation included 805 individuals. A total of 29 parameters were extracted from resting recordings of non-invasive continuous blood pressure recordings and high-resolution electrocardiograms of each subject. Gaussian process regression was then used to estimate effects of age on the cardiovascular system (see Figure 1).

**Figure 1.**
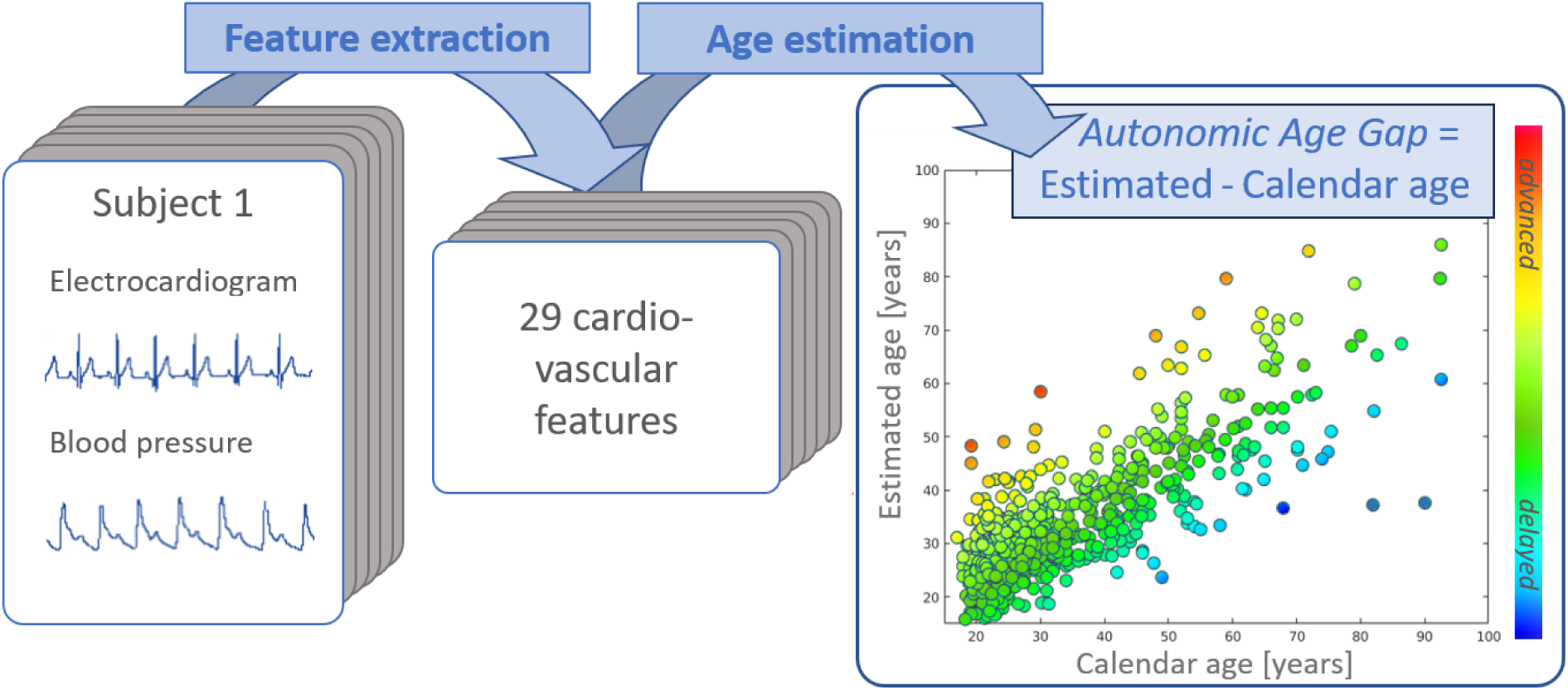
The framework for autonomic age gap estimation based on cardiovascular data. In total 29 indices were extracted from resting recordings of blood pressure and electrocardiograms. Machine learning (a Gaussian process regression model) was used to estimate age on the basis of these indices. The relation of estimated age and calendar age is depicted on the right side. The autonomic age gap is the difference between estimated and calendar age.

### Age estimation model and feature importance

In the validation sample, the model demonstrated a high concordance with chronological age, achieving a mean absolute error of MAE = 5.86 years and a root mean squared error of RMSE = 8.80 years. To identify the key contributors to age estimation, we calculated SHAP values (see Figure 2). The most important features were pulse reflection time, pulse transit time, and mean heart rate.

**Figure 2.**
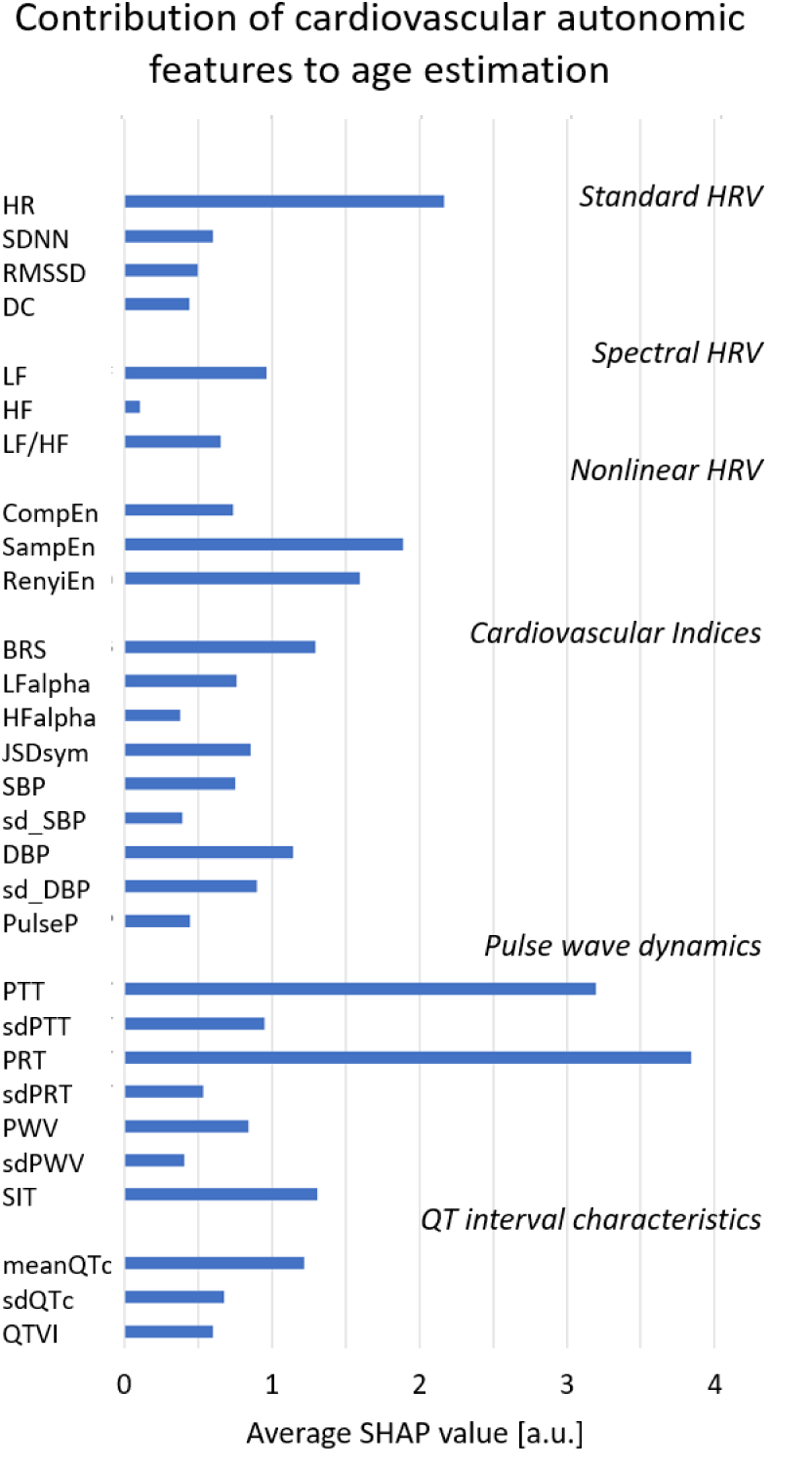
Averaged SHAP values of cardiovascular autonomic features that were fed into the age estimation model. HR: mean heart rate; SDNN: standard deviation of heart beat intervals (BBI); RMSSD: root mean square of successive BBI differences; DC: Deceleration capacity; LF: Low frequency spectral power of BBI; HF: High frequency spectral power of BBI; LF/HF: Low to high frequency spectral power ratio; CompEn: Compression entropy; SampEn: Sample entropy; RenyiEn: Renyi entropy; BRS: Baroreflex sensitivity; LFalpha: Low frequency cardiovascular coherence; HFalpha: High frequency cardiovascular coherence; JSDsym: Symmetric joint symbolic dynamics; SBP: Mean systolic blood pressure; sd_SBP: Standard deviation of systolic blood pressure; DBP: Mean diastolic blood pressure; sd_DBP: Standard deviation of diastolic blood pressure; PulseP: Pulse pressure; PTT: Mean pulse transit time; sdPTT: Standard deviation of PTT; PRT: Mean pulse rise time; sdPRT: Standard deviation of PRT; PWV: Mean pulse wave velocity; sdPWV: Standard deviation of PWV; SIT: Mean delay of dicrotic notch to pulse maximum; meanQTc: Mean corrected QT interval; sdQTc: Standard deviation of QT interval; QTVI: QT variability index.

### Age estimates in participants with low and high cardiovascular risk

Comparison of cardiovascular risk factors between the low-risk (Low CVR) and high-risk (High CVR) groups is summarized in Table 1. As expected, the Framingham risk score was significantly higher in the High CVR group, while normal risk estimates remained similar between the two groups, because these did not differ in age. The High CVR group had a higher prevalence of smokers and a higher BMI compared to the Low CVR group. In contrast, self-reported exercise levels and the presence of depressive symptoms did not differ significantly between the groups.

**Table 1.** Cardiovascular risk factors in the low-risk and high-risk group.

| Factor | Low CVR [N=61] | High CVR [N=72] | p-value |
| --- | --- | --- | --- |
| Age (years) | 52.5 ± 13.5 | 53.1 ± 12.9 | p=0.783 |
| Sex (m/w) | N=20/N=41 | N=31/N=41 | P=0.126 |
| Systolic blood pressure [mmHg] | 105.0 ± 10.4 | 133.0 ± 15.1 | p<0.001 |
| Body mass index [kg/m<sup>2</sup>] | 23.6 ± 2.8 | 26.8 ± 4.4 | p<0.001 |
| Smoker [yes/no] | N=0/N=61 | N=21/N=51 | p<0.001 |
| Framingham CVR [%] | 5.68 ± 5.27 | 12.10 ± 8.04 | p<0.001 |
| Framingham normal CVR [%] | 7.47 ± 5.87 | 7.68 ± 5.21 | p=0.827 |
| Regular exercise [yes/no] | N=46/N=15 | N=55/N=17 | p=0.895 |
| 1-2h/week | N=24 | N=27 |  |
| 3-4h/week | N=16 | N=22 |  |
| 5-6h/week | N=6 | N=6 |  |
| >6h/week | N=0 | N=0 |  |
| Depressive symptoms [BDI score] | N=4.88 ± 5.57 | 4.29 ± 4.44 | p=0.525 |

CVR: Cardiovascular risk, BDI: Beck's depression inventory.

The Gaussian process framework was used to estimate the a*utonomic age gap* (AAG) as a deviation from normal healthy aging. The model was trained on 809 individuals (324 men, 485 women, 31.8 ± 13.9 years) and validated on subset of 70 participants that was randomly selected and held out from training (26 men, 44 women, age: 30.7 ± 12.1 years). We applied the model to two groups: healthy participants with high cardiovascular risk (CVR) and those with low CVR. High CVR individuals had a significantly higher AAG (9.74 ± 17.78 years older) compared to the low CVR group (T = 3.91, p = 0.0004) and the validation set (T = 3.92, p = 0.0003), as shown in Figure 3. In contrast, age in the low CVR group was slightly underestimated (*AAG*: -2.22 ± 17.34 years). Predictions in the validation sample deviated minimally from calendar age, with an average error of 0.47 ± 8.85 years.

**Figure 3.**
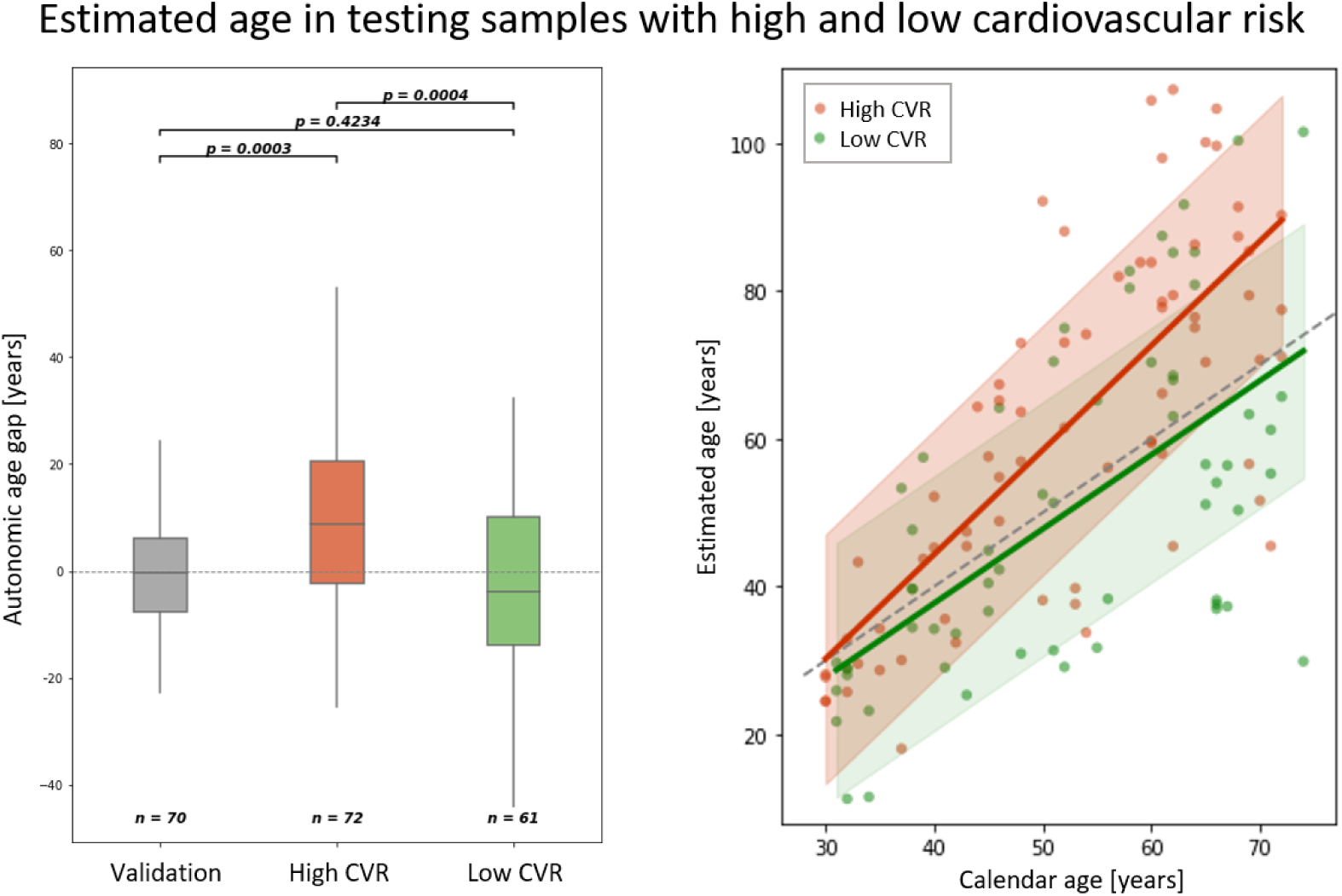
The autonomic age gap of participants with high cardiovascular risk (High CVR) and low cardiovascular risk (Low CVR). On average high-risk subjects showed increased autonomic age gap of 9.7 years (t-test, p<0.001). B: Estimated age in the High CVR group (red) and Low CVR group (green) as function of underlying calendar age. Shaded are indicate standard deviation of data points from linear fit.

A linear fit between cardiovascular-based age estimates and calendar age showed that the low CVR group had a near-ideal slope (m = 1.003) with a slight negative offset (intercept = -2.369 years). The high CVR group had a steeper slope (m = 1.41), but the difference between the two groups was not statistically significant (F = 3.16, p = 0.077).

## Discussion

Artificial intelligence has become an integral part of daily life, and machine learning (ML) is set to transform the medical and healthcare industries (39, 40). ML offers significant opportunities to enhance risk stratification, diagnostic classification, and patient subgroup identification, among other applications (41). One notable application of ML is the quantification of aging effects based on biological data, which has provided insights into the aging processes of different organ systems (42–44). Research suggests that these biological markers that track the aging process, the ‘aging clocks’, progress at individual rates which makes them a promising tool for precision medicine, particularly in addressing the diverse aging trajectories of older adults (3, 45).

In this study, we propose an ML-based framework to evaluate aging effects on cardiovascular autonomic regulation using a comprehensive set of biomarkers. *Autonomic age* quantifies the deviation between ML-predicted age and chronological age, providing insights into cardiovascular aging. We tested various ML algorithms to estimate age based on cardiovascular features (21), with the Gaussian Process Regression (GPR) model emerging as the most accurate approach in a large cohort of healthy individuals. Here, we assess the potential of this model to indicate cardiovascular risk by measuring deviations from the healthy aging trajectory.

Using deep learning, Attia et al. (2019) and Strodthoff et al. (2021) estimated chronological age based on short 12-lead ECG signals both achieving a MAE of 6.9 years. Attia et al (2019) reported that those patients, whose predicted age was more than 7 years higher than their calendar age, were more likely diagnosed with cardiovascular diseases such as hypertension and coronary disease. These studies highlight ML’s potential for cardiovascular risk assessment through age estimation.

Our model extends the range of input features for the model to learn to predict age by incorporating vascular indices, which are highly age-dependent. With aging, even in the absence of disease, large elastic arteries such as the aorta and carotid arteries become stiffer in both humans and animal models. Stiffening of arterial walls, containing baroreceptors, may compromise their sensory function, hindering the baroreflex mechanism (46). Additionally, age-related impairment of cardiovagal nerve traffic has been observed in older adults, potentially resulting in slower or attenuated heart rate responses (47). These mechanisms collectively contribute to impaired blood pressure regulation and an increased risk of sustained or orthostatic hypertension. Key indicators of age-related vascular changes include broad pulse waves, elevated pulse wave velocity, and increased systolic blood pressure.

Notably, the two most influential features in chronological age prediction were the pulse reflection time (PRT) and pulse transit time (PTT). Both parameters are closely linked to arterial stiffness, which progressively accelerates pulsatile blood flow with age (48). While these indicators were particularly relevant for the age estimation model, systolic blood pressure (SBP) had a comparatively lower impact, even though it is a widely used risk marker and contributed to cardiovascular risk (CVR) assessment in our study using the Framingham algorithm. Apart from SBP, none of the other input features were used for CVR assessment. However, the model still identified differences in cardiovascular aging between the two test sets due to their cardiovascular risk profile.

The average *autonomic age* indicated that healthy individuals with high CVR appeared from a cardiovascular perspective almost ten years older than expected. In contrast, the Low CVR group had a negative *AAG* of two years, suggesting advanced cardiovascular autonomic aging in those at increased risk for cardiovascular events. In the validation sample, *AAG* deviated by only half a year, reflecting a close alignment with chronological age. In the Low CVR group, the linear fit had a slope of approximately one, meaning the estimated age increased by one year for every additional year of chronological age. The slight negative offset may indicate better overall cardiovascular health persisting over a lifetime. Conversely, the High CVR group showed a steeper slope (1.4), suggesting accelerated aging processes—with estimated age increasing by 1.4 years for every additional year of chronological age.

The derived parameter *autonomic age* might serve as a comprehensive measure of autonomic status and as an independent risk marker that is easy to interpret. This is similar to ‘vascular age’ that has been described as clear and easy to understand by patients rather than an abstract mathematical construct (49). It has been demonstrated that using intelligible risk markers can motivate a population to adopt healthier lifestyles and improve CVD risk. Lopez-Gonzalez et al. (2015) have shown in a randomized clinical trial of 3,000 subjects that expressing cardiovascular risk in terms of years of age (‘heart age’) leads to decreases in risk scores including BMI reductions, compared to the use of traditional percentage-based markers.

Our study has several limitations that need to be noted. Although we investigated over a thousand subjects, especially at older ages, a rather small amount of data was available. However, recruitment of participants of an advanced age without suffering from cardiovascular, neurological or psychiatric disorder is very complicated. Furthermore, cognitive impairment, sensory loss, and changes in mobility might introduce a selection bias in our study. Finally, longitudinal studies are required in the future in order to track progression of *autonomic age* alongside chronological aging.

## Funding sources

This research was funded by the German Research Foundation (DFG, SCHU 3432/2-1) and the Interdisciplinary Center for Clinical Research Jena.

## Authorship contribution

AS conceived and designed research, performed experiments, analyzed data, prepared figures, interpreted results of experiments, drafted manuscript, edited and revised manuscript, approved final version of manuscript. YG analyzed data, prepared figures, drafted manuscript, approved final version of manuscript. DG interpreted results of experiments, edited and revised manuscript, approved final version of manuscript. ID interpreted results of experiments, edited and revised manuscript, approved final version of manuscript. MO interpreted results of experiments, edited and revised manuscript, approved final version of manuscript. CG: Interpreted results of experiments, edited and revised manuscript, approved final version of manuscript. KJB conceived and designed research, interpreted results of experiments, edited and revised manuscript, approved final version of manuscript

## Declaration of competing interest

The authors declare no conflict of interest.

## Data availability

The data used for training the model is publicly available at physionet.org (https://physionet.org/content/autonomic-aging-cardiovascular/1.0.0/)

## Notes

### Competing Interest Statement

The authors have declared no competing interest.

### Funding Statement

This study was funded by the German Research Foundation (DFG, SCHU 3432/2-1) and the Interdisciplinary Center for Clinical Research Jena.

### Author Declarations

Ethics committee of the Jena Univerity Hospital gave ethical approval for this work

